# Childhood growth and development and DNA methylation age in mid-life

**DOI:** 10.1101/2020.12.02.20242529

**Authors:** Jane Maddock, Juan Castillo-Fernandez, Andrew Wong, George B Ploubidis, Diana Kuh, Jordana T Bell, Rebecca Hardy

## Abstract

**Background:** In the first study of its kind, we examine the association between growth and development in early life and DNAm age biomarkers in mid-life.

**Methods:** Participants were from the Medical Research Council National Survey of Health and Development(n=1,376). Four DNAm age acceleration(AgeAccel) biomarkers were measured when participants were aged 53y:AgeAccelHannum, AgeAccelHorvath, AgeAccelLevine and AgeAccelGrim. Exposure variables included relative weight gain (standardised residuals from models of current weight z-score on current height, and previous weight and height z-scores) and linear growth (standardised residuals from models of current height z-score on previous height and weight z-scores) during infancy (0-2y, weight gain only), early childhood(2-4y), middle childhood(4-7y) and late childhood to adolescence(7- 15y), age at menarche and pubertal stage for men at 14-15y. The relationship between relative weight gain and linear growth and AgeAccel was investigated using conditional growth models. We replicated analyses from the late childhood to adolescence period and pubertal timing among 240 participants from The National Child and Development Study(NCDS).

**Results:** A 1 SD increase in relative weight gain in late childhood to adolescence was associated with 0.50y(95% CI:0.20,0.79) higher AgeAccelGrim. This was replicated in NCDS (0.57y(95%CI:-0.01, 1.16). A I SD increase in linear growth during early childhood was associated with lower AgeAccelLevine(−0.39y [95% CI:-0.74,-0.04) however we did not have the data to replicate this finding in NCDS. There was no strong evidence that relative weight gain and linear growth in childhood was associated with any other AgeAccel biomarker. There was no relationship between pubertal timing in men and AgeAccel biomarkers. Women who reached menarche ≥12y had 1.20y(95% CI:0.15,2.24) higher AgeAccelGrim on average than women who reached menarche <12y; however this was not replicated in NCDS.

**Conclusions:** Our findings generally do not support an association between growth and AgeAccel biomarkers in mid-life. However, rapid weight gain during pubertal development, which we found to be related to older AgeAccelGrim and had previously been related to higher cardiovascular disease risk, warrants further investigation.

## INTRODUCTION

The demographic shift towards an ageing population is a recognised public health challenge. Despite increases in life expectancy, compression of morbidity is not evident and there is significant heterogeneity in the occurrence of age-related disease and functional capability among people of the same chronological age^1^. Definitions of healthy ageing include survival to old age, delaying the onset of age-related diseases, and maintaining function^2^. Ageing is a complex process involving changes at the molecular, cellular, physiological and functional level over time^3^. Biomarkers of ageing, which combine information from one or more of these processes, have been proposed as tools to capture healthy ageing^4^. A suitable biomarker of ageing should be better at predicting survival, onset of age-related disease and functional capability at later ages than chronological age alone.

Epigenetic mechanisms, specifically DNA methylation (DNAm) have been implicated in the ageing process^5^. A number of DNAm-based biomarkers of ageing have been developed^6-11^. The first generation of these biomarkers used a data-driven elastic net regression method to identify specific DNAm sites (CpGs) that are highly predictive of chronological age. These DNAm-based biomarkers of ageing include the blood-based Hannum and the multi-tissue Horvath clocks^7,8^. The second generation of DNAm-based biomarkers of ageing, the Levine clock (also referred to as PhenoAge) and GrimAge, use information about age-related traits and mortality in addition to chronological age^10^. The Levine clock was developed using composite age-related clinical physiological measures to identify associated CpGs from DNAm in whole blood^10,11^. GrimAge was created by combining surrogate DNAm-based plasma protein estimates, DNAm smoking pack years estimates and chronological age and sex as a function of mortality ^11^. It remains unclear exactly what aspects of ageing each of these biomarkers are capturing^12^. However, having a higher DNAm age independent of chronological age (denoted age acceleration, AgeAccel) for each of these biomarkers has been shown to be associated with an increased risk of premature all-cause mortality, cardiovascular disease and cancer with AgeAccelLevine and AgeAccelGrim showing stronger associations than AgeAccelHannum or AgeAccelHorvath^10,11,13-16^. We have also recently demonstrated that AgeAccelLevine and AgeAccelGrim are associated with markers of age-related physical and cognitive performance^17^.

Using a life course approach can provide novel insights into how biological, behavioural and psychosocial processes over time affect healthy ageing^2^. Childhood is a sensitive period during which physiological changes can be initiated, leading to long-term health consequences in age-related physical and cognitive performance and age-related disease^18^. There may be a specific period during childhood where growth has a lasting impact on a particular age-related health outcome. Birth weight, child and adolescent weight and height gain have been associated with a range of age-related and disease outcomes but patterns vary depending on the outcome^19-24^. Similarly, the timing of puberty has exhibited differential associations with later life heath; for example younger age at puberty, particularly among women, is associated with higher risk of cardiovascular disease^25^ and all-cause mortality^26^ but later puberty is associated with lower bone mineral density^27^.

While a few studies in childhood and adolescence have examined how AgeAccelHorvath is associated with growth and pubertal timing (not vice-versa)^28-30^, to our knowledge, no study has examined how physical growth during childhood or pubertal timing is related to DNAm-based biomarkers of ageing in later life. In the largest of these previous studies (n=1018), higher AgeAccelHorvath at birth was associated with higher average fat mass and faster growth in weight and BMI between birth and 17 years^30^. In a cross-sectional study of Finnish children aged 11 to 13 years (n=239), higher AgeAccelHorvath was associated with heavier weight-for-age, taller height-for-age and more advanced puberty based on Tanner Stage^29^. In a smaller longitudinal (n=94) study of Chilean girls aged 9 to 13 years, higher AgeAccelHorvath was associated with earlier age at menarche^28^. These studies suggest that higher AgeAccelHorvath in early life is associated with more rapid growth and earlier development^28-30^. It is not known if this association is similar for other AgeAccel biomarkers, if it tracks across adulthood or if growth and development in childhood has additional effects on AgeAccel that persist across the life course.

In this exploratory study using data from a subsample of a nationally representative British birth cohort, we are the first to investigate the impact of birth weight and physical growth during infancy (birth to 2 years), early childhood (2 to 4 years), middle childhood (4 to 7 years) and late childhood to adolescence (7 to 15 years) and pubertal timing on four DNAm-based biomarkers of ageing in mid-life. We examine both linear growth and weight gain relative to linear growth to explore their potential separate effects^31^. Where possible we conducted a replication study among a sub-sample of participants from a British birth cohort born twelve years later.

## METHODS

### Participants

Participants were from the Medical Research Council National Survey of Health and Development study (MRC NSHD, or 1946 British birth cohort). NSHD is one of the longest running birth cohorts worldwide and participants have been followed up 24 times since birth. Details about this cohort have been published previously^32-34^. Briefly, the 5,362 original NSHD participants were singleton births born in one week in March 1946 to married parents in England, Scotland or Wales. While avoidable non-response in adulthood was highest among those with adverse socioeconomic circumstances and with low scores on childhood cognitive measures, study participants remain broadly representative of the native British population born in the early post-war years^34-36^. Of the 3,673 participants who were still alive and resident in England, Scotland or Wales when they were 53 years old, 3,035 provided information during a home visit by a research nurse and blood samples were taken from those who consented. The data collection at age 53 years received multicentre research ethics committee approval and informed consent was given by respondents to each set of questions and measures. A subset of participants at 53 years with blood samples who also had information on a wide range of health and age-related variables across the life course were selected for DNAm analyses (n=1,595). After quality control, 1,376 participants with DNAm information who also had weight and/or height measured at least once between birth and 15 years were included in analyses.

### DNAm-based biomarkers of ageing

DNAm from participant’s blood samples was measured at >850 000 CpG sites using Infinium MethylationEPIC BeadChips and processed using the ENmix package^37^ in R to obtain methylation beta-values for quality control purposes. In addition, signals with a detection p-value > 1×10-6 and a number of beads <3 were set to missing. Samples with missing data in >5% of the CpGs were excluded. CpGs with missing data in >5% of the samples were excluded. Samples identified with outlier values (more than three standard deviations from the mean or three interquartile ranges below the first or above the third quartiles) in bisulfite intensity, total intensity, or beta-value distribution were excluded. Sample identity was verified by estimating the correlation (r>0.90) between the 59 SNPs included in the methylation beadchips and imputed genotype data.

We used four DNAm-based biomarkers of ageing in this study: DNAm AgeHannum, DNAm AgeHorvath, DNAm AgeLevine and DNAm GrimAge^7,8,10,11^. DNAm GrimAge and DNAm AgeLevine were developed using both the Infinium HumanMethylation450 BeadChip and the Infinium MethylationEPIC BeadChips while DNAm AgeHannum and DNAm AgeHorvath used the Infinium HumanMethylation450 BeadChip only. Therefore participants included in this study have all CpGs for DNAm GrimAge and DNAm AgeLevine and are missing 6 CpGs for DNAm AgeHannum and 19 CpGs for DNAm AgeHorvath. Previous studies have found DNAm age estimate is unaffected by platform differences^38^. Besides those CpG sites already expected to be missing, in this study very few DNAm age CpG sites among a small number of participants did not pass quality control. Each DNAm-based biomarker was calculated using freely available software (https://labs.genetics.ucla.edu/horvath/dnamage/) with the normalisation option and advanced analysis for blood samples. Input data were produced using ssNoob pre-processing of the DNA methylation arrays in minfi^39^. Chronological age-independent DNAm-based biomarkers (residuals from a regression of DNAm age on chronological age) were calculated within this software to represent the difference between an individual’s DNAm Age and chronological age: AgeAccelHannum, AgeAccelHorvath, AgeAccelLevine and AgeAccelGrim (in units of a year). Estimated blood cell counts (naïve and exhausted CD8+ T-lymphocytes, CD4+ T-lymphocytes, B cells, natural killer cells, monocytes and granulocytes) were also calculated within this software.

### Weight and height

Weight (kg) and height (cm) in childhood were measured using standardised protocols at 2, 4, 7, 11 and 15 years. Birth weight was extracted from birth records to the nearest quarter pound and converted to kg. Birth length was not recorded. We conceptualised different periods of growth as: infancy (birth to 2 years), early childhood (2 to 4 years), middle childhood (4 to 7 years) and late childhood to adolescence (7 to 15 years which captures the full period of pubertal growth). All weight and height measures were converted to sex-specific z-scores using the mean and standard deviation to facilitate comparison of estimates between different ages.

### Pubertal timing

Pubertal stage for boys at age 14-15 years was based on physical examination by school doctors of four criteria: visibility of pigmented pubic hair, the visibility of axillary hair the development of genitalia, and whether the voice had broken ^40^. Based on responses to these questions boys were grouped into fully mature (i.e. those who experienced the earliest pubertal timing), advanced puberty, early puberty and pre-pubertal.

Age at menarche for girls was obtained from mother’s reports when the girls were 14-15 years. Of those who had not reached menarche at this date (*n*=188), age at menarche was obtained from self-report at 48 years (*n*=94).

Individual patterns of height and weight growth during puberty were estimated using the SITAR model of growth curve analysis and used in sensitivity analyses ^41^. Briefly, the SITAR model summarises each individual’s growth curve in terms of three parameters: size, tempo and velocity, each expressed relative to the mean curve. The tempo parameter indicates the relative timing of puberty based on the age at peak velocity with higher scores reflecting later pubertal timing.

### Additional variables

We selected a number of additional variables a priori for descriptive purposes. Socioeconomic position (SEP) at 53 years (or 43 years if missing) was based on occupation grouped according to the Registrar General’s social class and categorised into non-manual and manual. Similarly father’s SEP when the cohort member was 4 years old was categorised as non-manual or manual. Smoking status at 53 years was self-reported and categorised as current, ex-smoker or never smoker.

### Statistical analyses

All analyses were conducted in Stata 14 using the four AgeAccel biomarkers as outcomes. Further mention of AgeAccel refers to all four biomarkers unless specified. Models using AgeAccel as an outcome were adjusted for sex and age at home visit (in months) when DNA sample was taken.

We assessed if participants characteristics differed between those included in our main analyses and those who responded to the 53 year data collection but were not included in our analyses using t-tests and chi-squared tests.

In preliminary analyses we investigated associations between weight and height from infancy to adolescence and AgeAccel. The relation of weight and height z-scores at each age with AgeAccel was examined using separate multiple regression models for each age for descriptive purposes. Models were mutually adjusted for weight or height z-scores. Interactions between sex and height or weight z-scores were tested to assess whether associations were different for men and women.

For our main analyses, we used regression with conditional growth measures ^42-45^. This method involves computing a sex-specific conditional growth measure i.e. the standardised residuals from a regression of current size on the previous size measure. These conditional growth variables are, by definition, uncorrelated with size at the previous age and represent the deviation of a participant’s current size from that expected given their previous measure and the growth of the other participants in the sample. In order to examine the effects of both linear growth and relative weight gain, we used an approach described by Adair and colleagues ^31^. Conditional relative weight was calculated as the standardised residuals of current weight z-score accounting for previous weight and height z-score as well as current height z-score. Conditional height was calculated as the standardised residuals from a regression of current height z-score on previous height and weight z-scores (but not current weight z-score). For each period of childhood, linear regression models were used to examine the association between the conditional growth measure and AgeAccel.

Linear regression models were also used to examine the association between pubertal timing and AgeAccel. For girls, we used both continuous age at menarche and a dichotomised score of <12 years and ≥ 12 years to examine early menarche. For boys pubertal stage at 14-15 years was assessed.

#### Sensitivity analyses

Since more participants had information on the SITAR variables than timing of puberty, we repeated the analysis using height-tempo.

We adjusted the conditional growth models and timing of puberty for estimated cell counts (naïve and exhausted CD8+ T-lymphocytes, CD4+ T-lymphocytes, B cells, natural killer cells, monocytes and granulocytes).

A subsample of participants in NSHD also had AgeAccel measures at 60-64 years (n=482). Therefore we investigated if the association between growth and development in early life and AgeAccel were consistent when AgeAccel was measured approximately ten years later.

### Replication

We used data from the National Child and Development Study (NCDS; 1958 British birth cohort) to replicate findings where possible. This cohort has been described in detail previously ^46^. For this analysis we used data from a subsample of NCDS participants who had DNAm measured at 45 years (n=240). AgeAccel measures were obtained using the same methods as NSHD. Height and weight were measured at ages 7, 11 and 16 years. We used weight z-scores in NCDS at 7 and 16 years to replicate analyses for growth during the late childhood to adolescence period and AgeAccel observed in NSHD using regression with conditional growth measures as outlined above.

Pubertal stage among boys in NCDS was assessed by a physical examination by trained medical personnel when participants were 16 years. We used the following criteria to assign boys as fully mature (i.e. those who experienced the earliest pubertal timing) or later puberty for comparison with NSHD: visibility of pigmented pubic hair, the visibility of axillary hair, whether the voice had broken and the visibility of facial hair. Age at menarche was reported by the medical officer (or parent if missing) at the 16 year examination. Two women in the NCDS sample had not reached menarche by 16 years and were coded as greater than 16 years.

## RESULTS

Descriptive characteristics of participants included in the main analysis (n=1,376) are outlined in table 1. There were no major differences in body size, pubertal timing, smoking status or SEP among participants included in our main analysis versus all other NSHD participants responding to the 53 year data collection (n=1,659, supplementary table 1).

**Table 1.** Descriptive characteristics (n=1376)

| **Table 1.** Descriptive characteristics (n=1376) |  |  |
| --- | --- | --- |
|  | Men (n=656) | Women (n=720) |
| AgeAccel measures at 53 years, mean (SD) |  |  |
| DNAm age Hannum (y) | 43.1 (4.3) | 41.6 (4.0) |
| AgeAccelHannum | 0.79 (4.28) | -0.66 (3.95) |
| DNAm age Horvath (y) | 50.7 (4.2) | 49.6 (3.9) |
| AgeAccelHorvath | 0.54 (4.15) | -0.54 (3.86) |
| Levine (y) | 39.0 (5.6) | 38.9 (5.6) |
| AgeAccelLevine | 0.06 (5.59) | -0.02 (5.61) |
| GrimAge (y) | 58.0 (5.1) | 55.3 (4.8) |
| AgeAccelGrim | 1.40 (5.14) | -1.28 (4.79) |
| Weight (kg), mean (SD) |  |  |
| At birth | 3.46 (0.53) | 3.34 (0.49) |
| At 2 years | 13.23 (1.50) | 12.54 (1.39) |
| At 4 years | 17.42 (2.10) | 16.84 (2.02) |
| At 7 years | 23.00 (2.84) | 22.39 (3.11) |
| At 15 years | 51.83 (9.73) | 51.53 (8.58) |
| Height (cm), mean (SD) |  |  |
| At 2 years | 86.11 (5.08) | 84.81 (4.44) |
| At 4 years | 103.47 (5.15) | 102.54 (4.87) |
| At 7 years | 120.49 (5.86) | 119.42 (5.36) |
| At 15 years | 162.24 (9.17) | 158.51 (6.05) |
| Pubertal Stage at 14-15 years (men), % (N) |  |  |
| Fully mature* | 26.22 (172) |  |
| Advanced puberty | 32.16 (211) |  |
| Early puberty | 30.49 (200) |  |
| Pre-pubertal | 11.13 (73) |  |
| Age at menarche (y) (women), mean (SD) |  | 13.11 (1.26) |
| *Fully mature group are the group who experienced the earliest pubertal timing<br>AgeAccel: Age Acceleration |  |  |

The median absolute difference between DNAm AgeHannum, DNAm AgeHorvath, DNAm Levine and DNAm GrimAge and chronological age at 53 years was 11.1, 4.0, 14.7 and 2.8 years respectively. The correlation coefficients between the different AgeAccel biomarkers ranged from *r*=0.1 for AgeAccelHorvath and AgeAccelGrim to *r*=0.4 for AgeAccelLevine with AgeAccelHannum, AgeAccelHorvath and AgeAccelGrim.

### Preliminary analyses: AgeAccel at 53 years and weight and height z-scores at each time point in childhood

We find little evidence of any strong relationships between weight and height at each age separately with any AgeAccel biomarker at 53 years (figure 1). The largest differences in coefficients from one age to the next, which are indicative of the importance of change in size, are observed between 7 and 15 years for weight in relation to AgeAccelGrim and also with AgeAccelHannum and AgeAccelLevine. There were also differences in coefficients observed between 2 and 4 years and 4 and 7 years for height in relation to AgeAccelHanum and AgeAccelLevine. There was no consistent evidence for sex differences except for AgeAccelHorvath where there was evidence for an interaction between sex and birth weight z-score and weight z-score at 4 years (p_interaction_≤0.03).

**Figure 1.**
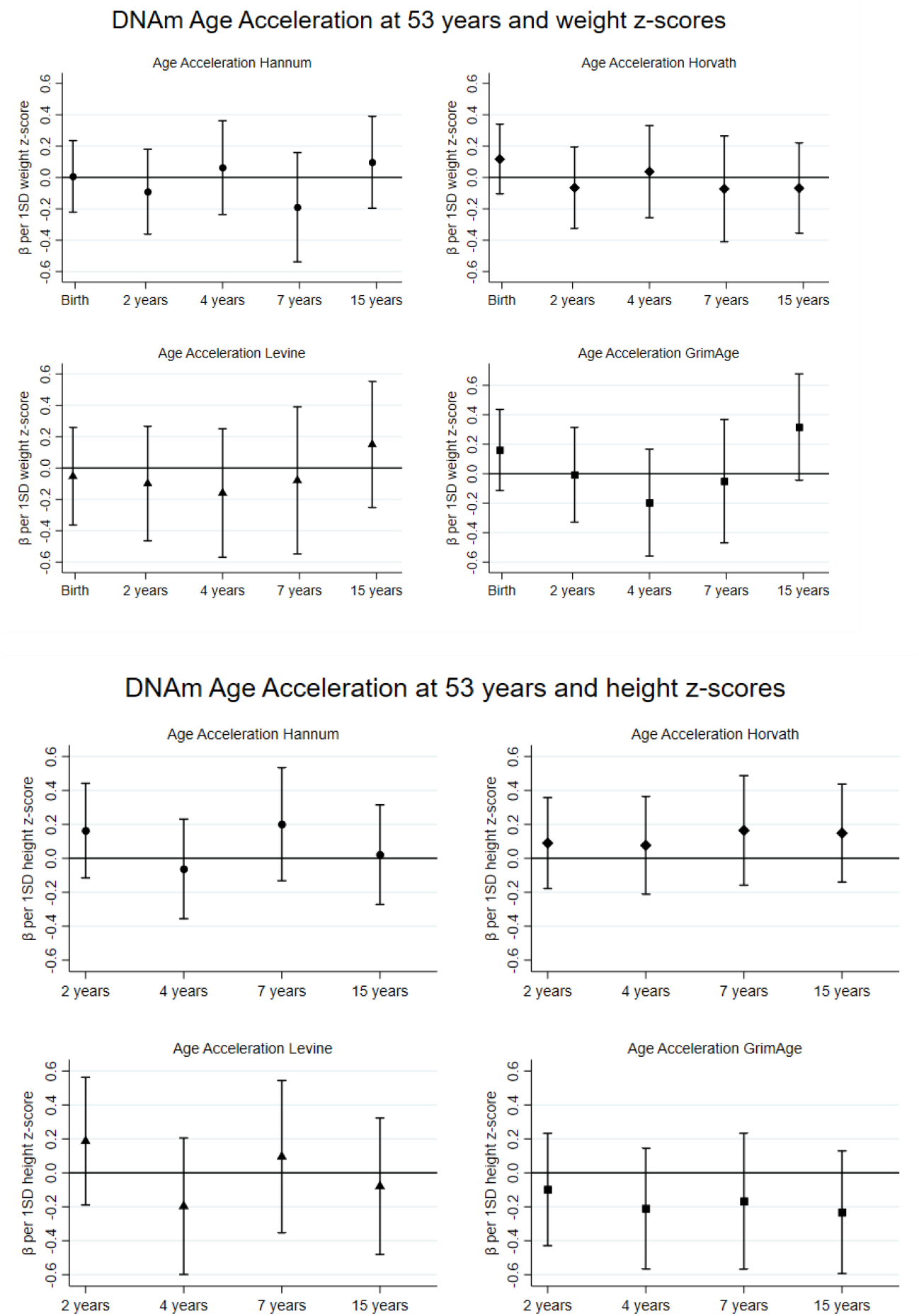
Preliminary results for DNAm Age Acceleration at 53 years and weight and height z-scores. Adjusted for height/weight z-score, age in months at 53 years and sex. Each coefficient represents mean change in AgeAccel (y) for a 1 SD increase in height/weight. Separate analyses were conducted at each age.

### Main analyses: AgeAccel at 53 years and conditional relative weight and linear growth in childhood

There was no evidence that relative weight gain and linear growth during childhood was associated with AgeAccelHannum or AgeAccelHorvath (table 2).

**Table 2.** DNAm Age Acceleration at 53 years and conditional growth.

| **Table 2.** DNAm Age Acceleration at 53 years and conditional growth |  |  |  |  |  |  |  |  |  |
| --- | --- | --- | --- | --- | --- | --- | --- | --- | --- |
|  |  | AgeAccelHannum |  | AgeAccelHorvath |  | AgeAccelLevine |  | AgeAccelGrim |  |
|  | N | Coefficient (95% CI) | P value | Coefficient (95% CI) | P value | Coefficient (95% CI) | P value | Coefficient (95% CI) | P value |
| *Relative weight gain* |  |  |  |  |  |  |  |  |  |
| RWG between birth and 2 years | 1,127 | -0.09 (-0.34, 0.16) | 0.48 | -0.09 (-0.34, 0.15) | 0.44 | -0.05 (-0.39, 0.29) | 0.76 | -0.08 (-0.38, 0.22) | 0.62 |
| RWG between 2 and 4 years | 1,065 | 0.11 (-0.15, 0.37) | 0.41 | 0.18 (-0.07, 0.43) | 0.16 | -0.07 (-0.42, 0.28) | 0.69 | -0.11 (-0.42, 0.19) | 0.47 |
| RWG between 4 and 7 years | 1,168 | -0.17 (-0.43, 0.08) | 0.18 | -0.08 (-0.33, 0.16) | 0.51 | 0.05 (-0.29, 0.40) | 0.76 | 0.08 (-0.23, 0.39) | 0.60 |
| RWG between 7 and 15 years | 1,161 | 0.16 (-0.08, 0.40) | 0.19 | -0.01 (-0.24, 0.23) | 0.94 | 0.22 (-0.11, 0.54) | 0.19 | 0.50 (0.20, 0.79) | <0.001 |
| *Linear growth* |  |  |  |  |  |  |  |  |  |
| CLG between 2 and 4 years | 1,085 | -0.06 (-0.31, 0.20) | 0.67 | 0.07 (-0.18, 0.32) | 0.57 | -0.39 (-0.74, -0.04) | 0.03 | -0.24 (-0.55, 0.06) | 0.12 |
| CLG between 4 and 7 years | 1,204 | 0.11 (-0.12, 0.33) | 0.36 | -0.04 (-0.26, 0.19) | 0.75 | 0.16 (-0.15, 0.47) | 0.32 | 0.01 (-0.27, 0.28) | 0.97 |
| CLG between 7 and 15 years | 1,174 | 0.11 (-0.13, 0.35) | 0.36 | 0.03 (-0.20, 0.26) | 0.78 | 0.01 (-0.31, 0.33) | 0.97 | 0.14 (-0.15, 0.43) | 0.35 |
| Adjusted for age in months at 53 years and sex. |  |  |  |  |  |  |  |  |  |
| RWG: Relative weight gain i.e. standardised residuals from regression of present weight z-score on previous weight and height z-scores and present height z-score |  |  |  |  |  |  |  |  |  |
| CLG: Conditional linear growth i.e. Standardised residuals from regression of present height z-score on previous height and weight z-scores |  |  |  |  |  |  |  |  |  |

In the conditional growth models (table 2) a 1 SD increase in relative weight gain between the ages of 7 and 15 years was associated with 0.50 years (95% CI: 0.20, 0.79) higher AgeAccelGrim.

For linear growth, there was modest evidence that more rapid growth between 2 and 4 years was associated with lower AgeAccelLevine (−0.39 years [95% CI: −0.74, −0.04]).

### Main analyses: AgeAccel at 53 years and pubertal timing

There was no relationship between pubertal timing in men and any of the AgeAccel biomarkers at 53 years (table 3). Women who reached menarche at 12 years or older had 1.20 years (95% CI: 0.15, 2.24) higher AgeAccelGrim on average than women who reached menarche younger than 12 years.

**Table 3.** DNAm Age Acceleration at 53 years and pubertal timing.

| **Table 3. DNAm Age Acceleration at 53 years and pubertal timing** |  |  |  |  |  |  |  |  |  |
| --- | --- | --- | --- | --- | --- | --- | --- | --- | --- |
|  |  | AgeAccelHannum |  | AgeAccelHorvath |  | AgeAccelLevine |  | AgeAccelGrim |  |
|  | N | Coefficient (95% CI) | P value | Coefficient (95% CI) | P value | Coefficient (95% CI) | P value | Coefficient (95% CI) | P value |
| *Women* |  |  |  |  |  |  |  |  |  |
| Age at menarche (years) | 617 | 0.07 (-0.18, 0.32) | 0.59 | 0.04 (-0.20, 0.29) | 0.73 | -0.02 (-0.37, 0.33) | 0.91 | 0.22 (-0.08, 0.53) | 0.15 |
| Age at menarche |  |  |  |  |  |  |  |  |  |
| <12 years | 96 | Ref |  | Ref |  | Ref |  | Ref |  |
| ≥12 years | 521 | 0.37 (-0.49, 1.23) | 0.40 | 0.27 (-0.57, 1.11) | 0.53 | 0.73 (-0.48, 1.94) | 0.24 | 1.20 (0.15, 2.24) | 0.03 |
| *Men: Pubertal status at 14-15 years* |  |  |  |  |  |  |  |  |  |
| Fully mature* | 172 | Ref. |  | Ref. |  | Ref. |  | Ref. |  |
| Advanced puberty | 211 | 0.17 (-0.70, 1.03) | 0.56** | -0.36 (-1.20, 0.48) | 0.83** | 0.95 (-0.17, 2.08) | 0.08** | -0.29 (-1.33, 0.75) | 0.93** |
| Early puberty | 200 | 0.39 (-0.48, 1.27) |  | -0.18 (-1.03, 0.66) |  | 0.50 (-0.64, 1.64) |  | -0.05 (-1.10, 1.00) |  |
| Pre-pubertal | 73 | -0.40 (-1.57, 0.77) |  | -0.39 (-1.54, 0.75) |  | -0.83 (-2.36, 0.70) |  | -0.30 (-1.71, 1.11) |  |
| Fully mature | 172 | Ref. |  | Ref. |  | Ref. |  | Ref. |  |
| Later puberty | 484 | 0.18 (-0.57, 0.92) | 0.64 | -0.29 (-1.02, 0.43) | 0.43 | 0.50 (-0.48, 1.47) | 0.32 | -0.19 (-1.09, 0.70) | 0.67 |
| Adjusted for age in months at 53 years. *Fully mature group are the group who experienced the earliest pubertal timing. **p-value from lrtest comparing models with and without categorical puberty variable |  |  |  |  |  |  |  |  |  |

### Sensitivity analyses

We observed no associations between the SITAR measure of pubertal timing in men or women with any AgeAccel biomarker (supplementary table 2).

Adjusting the conditional growth models for estimated cell composition attenuated the estimates; the estimate for the association of linear growth between 2-4 years and AgeAccelLevine was halved to −0.20 (95% CI: −0.50 to 0.10) (supplementary table 3). The association between age at menarche and AgeAccelGrim was greatly attenuated to 0.53 years (95% CI: −0.46, 1.53) when adjusting for estimated cell composition (supplementary table 4).

When using AgeAccel at 60-64 years as the outcome where the sample size is smaller, the association between relative weight gain between 7 and 15 years and AgeAccelGrim was weaker than that observed at 53 years (0.38 [95% CI: −0.04 to 0.79] (supplementary table 5). An association between relative weight gain between 7 and 15 years and AgeAccelLevine was observed at 60-64 years (0.69 [95% CI: 0.12, 1.26]. The estimated association between linear growth between 2 and 4 years and AgeAccelLevine was similar (−0.37 [95% CI: −0.97 to 0.22]) to that found at age 53 years.

The direction of the estimates between pubertal timing and AgeAccel using the subsample from NSHD at 60-64 years were in a similar direction as 53 years. Associations between age at menarche (≥12 years versus <12 years) and AgeAccelLevine at 60-64 years were stronger compared with AgeAccel biomarkers from 53 years (supplementary table 6). Among men, much larger estimates were observed when compared with 53 years. For example, compared to men with early puberty, those with later puberty had 1.27 years lower (95% CI: −2.47, - 0.06) AgeAccelGrim at 60-64 years and 0.19 years lower (95% CI: −1.09, 0.70) at 53 years.

### Replication in NCDS

Similar to results using NSHD participants at 53 years, a 1 SD increase in relative weight gain between the ages of 7 and 16 years was associated with 0.20 (95% CI −0.47 to 0.86) years higher AgeAccelGrim (table 4). There was no association between pubertal timing and any AgeAccel biomarker in NCDS.

**Table 4.** DNAm Age Acceleration at 45 years, conditional growth between 7 and 16 years and pubertal timing in the National Child and Development Study.

|  |  |  |  |  |  |  |  |  |  |
| --- | --- | --- | --- | --- | --- | --- | --- | --- | --- |
|  | N | AgeAccelHannum |  | AgeAccelHorvath |  | AgeAccelLevine |  | AgeAccelGrim |  |
|  |  | Coefficient (95% CI) | P value | Coefficient (95% CI) | P value | Coefficient (95% CI) | P value | Coefficient (95% CI) | P value |
| **Conditional growth** |  |  |  |  |  |  |  |  |  |
| RWG between 7 and 16 years | 240 | -0.04 (-0.48, 0.39) | 0.84 | 0.23 (-0.25, 0.70) | 0.35 | 0.20 (-0.47, 0.86) | 0.56 | 0.57 (-0.01, 1.16) | 0.06 |
| **Pubertal timing** |  |  |  |  |  |  |  |  |  |
| *Women* |  |  |  |  |  |  |  |  |  |
| Age at menarche (years)* | 112 | -0.21 (-0.76, 0.33) | 0.44 | -0.20 (-0.77, 0.38) | 0.50 | -0.21 (-0.94, 0.53) | 0.58 | -0.08 (-0.71, 0.55) | 0.80 |
| Age at menarche |  |  |  |  |  |  |  |  |  |
| <12 years | 12 | Ref |  | Ref |  | Ref |  | Ref |  |
| ≥12 years | 100 | -0.77 (-3.02, 1.47) | 0.50 | -0.71 (-3.07, 1.65) | 0.55 | -1.06 (-4.07, 1.96) | 0.49 | -0.01 (-2.61, 2.58) | 0.99 |
| *Men: Pubertal status at 16 years* |  |  |  |  |  |  |  |  |  |
| Fully mature** | 13 | Ref. |  | Ref. |  | Ref. |  | Ref. |  |
| Later puberty | 99 | 0.08 (-1.77, 1.93) | 0.93 | -0.002 (-2.050, 2.047) | 1.00 | -0.99 (-4.29, 2.31) | 0.55 | -1.50 (-4.38, 1.39) | 0.31 |
| Adjusted for sex. RWG: Relative weight gain i.e. standardised residuals from regression of weight z-score at 16 years on weight and height z-scores at 7 years and height z-score at 16 years. CLG: Conditional linear growth i.e. Standardised residuals from regression of height z-score at 16 years on height and weight z-scores at 7 years *Those starting after 16 years coded as ≥16 years **Fully mature group are the group who experienced the earliest pubertal timing |  |  |  |  |  |  |  |  |  |

## DISCUSSON

We did not find strong evidence for an association between growth in early life and all AgeAccel biomarkers in mid-adulthood. However, we did observe an association between faster weight gain during pubertal growth and higher AgeAccelGrim in mid-adulthood which was seen in NSHD and replicated in NCDS.

There are no previous studies with which to directly compare our findings. However in an English study, higher birth weight was correlated with higher AgeAccelHorvath at 7 years but with lower AgeAccelHorvath by 17 years, with no information for correlations beyond adolescence ^47^. In the same study looking at the association in the other direction, higher AgeAccelHorvath at birth was associated with more rapid childhood and adolescent development including faster weight and BMI gains between childhood and adolescence ^30^. Similarly, in a cross-sectional study of Finnish children aged 11 to 13 years (n=239), higher AgeAccelHorvath was associated with heavier weight-for-age and taller height-for-age ^29^. We did not observe any strong associations with birth weight and AgeAccelHorvath in mid- life suggesting that if these associations do exist, they may not persist into adulthood.

We did observe a relationship between faster gains in weight during pubertal growth with AgeAccelGrim. Among adults there is a consistent association between higher BMI and higher AgeAccel in all four biomarkers ^10,11,48^. Since rapid pubertal weight gain is associated with higher adult BMI, our findings suggest that the relationship we observed with AgeAccelGrim in mid-life may have been at least partly established in early life.

Our finding of an association between faster linear growth in early childhood and higher AgeAccelLevine among NSHD participants should be interpreted with caution. The estimates we observed for the association between faster linear growth in early childhood and higher AgeAccelLevine were not as large as the estimates for rapid weight gain in adolescence and multiple tests were conducted. Adjusting for cell composition attenuated the association observed between linear growth and AgeAccelLevine however, since cell composition is a component in the creation of DNAm age Levine, this could be an over adjustment. We were unable to replicate this finding due to a lack of early life growth measures in NCDS.

While two previous studies in Finland and Chile observed that higher AgeAccelHorvath among children aged 9 to 13 years was associated with more advanced puberty based on Tanner stage or earlier age at menarche^28,29^, to our knowledge no previous study has examined the relationship between pubertal timing and AgeAccel biomarkers in later life. We found no evidence for an association between pubertal timing in men and any of the AgeAccel biomarkers at 53 years in NSHD or at 45 years in NCDS. Early puberty was associated lower AgeAccelGrim at 60-64 years among men in NSHD, however this was a small sample and would need replication in a larger study. An association between older age at menarche and higher AgeAccel was observed in NSHD at both 53y and 60-64 years. This finding is unexpected as older age at menarche is generally associated with better age-related outcomes and reduced risk of mortality ^26^. This association was not replicated in NCDS or when using age at menarche as a continuous variable. We also observed no association when SITAR variables were used to represent pubertal timing. Previous analysis in NSHD found no association between age at menarche and all-cause mortality suggesting that different mortality rates by pubertal timing in women are unlikely to have biased this finding ^49^. In sensitivity analyses we found that that the associations between older age at menarche and higher AgeAccel attenuated following adjustment for cell composition. As there is some evidence that pubertal timing is associated with white blood cell counts ^50^, it is possible that blood cell counts confounded our observed association. With all this in mind, these associations should be interpreted with caution and require further investigation in larger samples.

The use of DNAm to predict biological age is a newly emerging field and there are still lot of unknowns. It is difficult to ascertain what aspects of ageing these biomarkers are capturing and the underlying biology of these relationships ^12^. For this reason we examined four AgeAccel biomarkers with the aim of informing future studies. We observed weak correlations between the four AgeAccel biomarkers suggesting that they are not necessarily capturing the same underlying ageing construct. There are a few differences between how these biomarkers were constructed that may explain the differences in the results. The first generation of biomarkers are generally considered chronological age predictors and the models were trained (i.e. the CpG sites and weights were determined) primarily using cross-sectional data. The second generation biomarkers used additional age-related outcomes to select the CpG sites and were trained using longitudinal data ^12^. The nature of the relationships in our study may provide more insight into the utility of these biomarkers.

Assuming that growth in early life has an effect on age-related health outcomes, our finding of no associations between growth from infancy to middle childhood in any of the AgeAccel biomarkers suggest that these biomarkers are not fully capturing the effects of early life growth on age-related conditions. However, the finding of an association between faster gains in weight during adolescence and higher AgeAccelGrim and a suggested association between faster linear growth during childhood with higher AgeAccelLevine is similar to that seen with cardiovascular risk factors such as obesity, blood pressure and vascular structure and cardiovascular disease ^19-21,51^. This may suggest that the second generation AgeAccel biomarkers (particularly AgeAccelGrim) are capturing the result of a cardiovascular pathway through which pubertal growth influences age-related disease, perhaps due to the inclusion of cardiovascular risk-factors in their construction.

Our main results are based on relatively young participants at 53 years. While age-related disease may not always be evident at this age, it is possible that age-related DNAm changes are occurring. In order to examine if our observed associations change with age we repeated analyses on a subsample of NSHD participants who also had AgeAccel measures ten years later. We found that the estimates at 60-64 years were generally in the same direction but that some associations were weaker and some were stronger at 60-64 years. As participants age and accumulate more age-related changes the effect of early life development may become more evident. However, given the small sample size at 60-64 years, these findings would need replication in larger studies.

The main strengths of this study are the inclusion of participants from two well characterised prospective population-based birth cohorts, the prospective repeated measures of body size from infancy, and the replication of the main finding. There are also a number of limitations to keep in mind when interpreting these findings. We conducted 44 tests for our main analyses. As this was an exploratory study, we decided not to adjust for multiple testing ^52,53^. Therefore, our finding of an association between relative weight gain during pubertal growth and AgeAccelGrim could be by chance and although this was replicated in another sample, future replication is required before any conclusions are drawn. As with all prospective cohort studies there is attrition in NSHD; however at 53 years respondents were found to be generally representative of the white British population ^32^. We observed no major differences in sociodemographic characteristics between participants included in our analyses and those who responded to the 53 year data collection but were not included in our analyses. There is a possibility that collider bias could have been introduced, as the selection for having DNAm was from those with complete growth and development data in childhood. If having lower DNAm age and faster weight gain was associated with participation, the estimates may have been biased ^54^. Where possible we repeated our analyses in a subsample from NCDS to replicate findings, which could be similarly biased. Since measures were not obtained from exactly the same ages we were unable to test for replication of the associations in infancy, early childhood or middle childhood. Similarly, timing of puberty was assessed slightly differently between the studies which may have accounted for some differences in findings. Finally, as with all observational studies, unmeasured cofounding remains a limitation. We decided a priori to adjust only for sex and age at home visit. A thorough investigation of potential confounders and/or mediators would be required before any inference to causality could be made.

Our study serves two purposes: examining the utility of newly emerging aging biomarkers, and the importance of growth and development in early life on ageing. Our findings suggest that in general these AgeAccel biomarkers do not capture the age-related effects of childhood growth. The second generation AgeAccel biomarkers, particularly AgeAccelGrim appear to be more sensitive to growth during puberty. The observed relationship between faster gains in weight during puberty, which have previously been associated with cardiovascular risk, and AgeAccelGrim indicates that this period of growth warrants further investigation.

## Data Availability

Data used in this publication are available to bona fide researchers upon request to the NSHD Data Sharing Committee via a standard application procedure. Further details can be found at http://www.nshd.mrc.ac.uk/data doi: 10.5522/NSHD/S201. Information about NCDS can be found at https://cls.ucl.ac.uk/cls-studies/1958-national-child-development-study/ and data can be accessed via https://ukdataservice.ac.uk/.

## Acknowledgments

We thank NSHD and NCDS study members for their lifelong participation and past and present members of the study teams, including Professor Alissa Goodman, Professor Ken Ong and members of the MRC Epidemiology unit in Cambridge, who helped to collect and process the data. Data used in this publication are available to bona fide researchers upon request to the NSHD Data Sharing Committee via a standard application procedure. Further details can be found at http://www.nshd.mrc.ac.uk/data doi: 10.5522/NSHD/S201. Information about NCDS can be found at https://cls.ucl.ac.uk/cls-studies/1958-national-child-development-study/ and data can be accessed via https://ukdataservice.ac.uk/.

