## Supplementary Materials for "Childhood growth and development and DNA methylation age in mid-life"

**Supplementary table 1**. Comparison of participants included in this analytical sample versus all other participants responding to 53 year data collection

| | Included (n=1376) | | | Not included (n=1,659) | | |
|---|---|---|---|---|---|---|
| | n | | | n | | P-value |
| *Weight (kg), mean (SD)* | | | | | | |
| At birth | 1,375 | 3.40 (0.51) | | 1,650 | 3.39 (0.50) | 0.77 |
| At 2 years | 1,203 | 12.88 (1.49) | | 1,331 | 12.95 (1.52) | 0.22 |
| At 4 years | 1,298 | 17.12 (2.08) | | 1,423 | 17.37 (2.20) | 0.003 |
| At 7 years | 1,299 | 22.70 (2.99) | | 1,223 | 22.91 (3.14) | 0.08 |
| At 15 years | 1,232 | 51.68 (9.18) | | 1,113 | 51.98 (8.52) | 0.42 |
| *Height (cm) , mean (SD)* | | | | | | |
| At 2 years | 1,174 | 85.45 (4.81) | | 1,296 | 85.18 (5.11) | 0.19 |
| At 4 years | 1,264 | 102.99 (5.02) | | 1,400 | 103.35 (5.11) | 0.06 |
| At 7 years | 1,334 | 119.95 (5.64) | | 1,280 | 120.07 (5.56) | 0.58 |
| At 15 years | 1,234 | 160.39 (8.00) | | 1,116 | 160.25 (7.63) | 0.65 |
| *Pubertal Stage at 14-15 years (men), %* | | | | | | |
| Fully mature | 172 | 26.2 | | 125 | 22.9 | 0.19 |
| Later puberty | 484 | 73.9 | | 420 | 77.1 | |
| *Age at menarche (y) (women) , mean (SD)* | 617 | 13.1 (1.3) | | 610 | 13.0 (1.2) | 0.11 |
| *Socioeconomic position (%)* | | | | | | |
| Non-manual | 891 | 66.79 | | 1,042 | 66.79 | 0.99 |
| Manual | 443 | 33.21 | | 518 | 33.21 | |
| *Smoking status* | | | | | | |
| Current smoker | 333 | 24.22 | | 362 | 22.44 | 0.24 |
| Ex-smoker | 630 | 45.82 | | 788 | 48.85 | |
| Never smoker | 412 | 29.96 | | 463 | 28.70 | |

*Fully mature group are the group who experienced the earliest pubertal timing

**Supplementary table 2**. DNAm Age Acceleration at 53 years and SITAR height-tempo

| | | AgeAccelHannum | | AgeAccelHorvath | | AgeAccelLevine | | AgeAccelGrim | |
|---|---|---|---|---|---|---|---|---|---|
| | N | Coefficient (95% CI) | P value | Coefficient (95% CI) | P value | Coefficient (95% CI) | P value | Coefficient (95% CI) | P value |
| *Women* | | | | | | | | | |
| Height tempo (years) | 720 | 0.01 (-0.03, 0.06) | 0.49 | -0.02 (-0.06, 0.02) | 0.35 | -0.02 (-0.08, 0.03) | 0.42 | 0.02 (-0.02, 0.07) | 0.33 |
| *Men* | | | | | | | | | |
| Height tempo (years) | 656 | -0.050 (-0.101, 0.002) | 0.06 | -0.02 (-0.07, 0.03) | 0.38 | -0.04 (-0.10, 0.03) | 0.30 | -0.001 (-0.063, 0.061) | 0.97 |

Adjusted for age at interview (in months) at 53 years

**Supplementary table 3.** DNAm Age Acceleration at 53 years and conditional growth adjusted for estimated cell composition

| | | AgeAccelHannum | | AgeAccelHorvath | | AgeAccelLevine | | AgeAccelGrim | |
|---|---|---|---|---|---|---|---|---|---|
| | N | Coefficient (95% CI) | P value | Coefficient (95% CI) | P value | Coefficient (95% CI) | P value | Coefficient (95% CI) | P value |
| *Relative weight gain* | | | | | | | | | |
| RWG between birth and 2 years | 1,127 | -0.12 (-0.34, 0.10) | 0.29 | -0.09 (-0.33, 0.14) | 0.44 | -0.11 (-0.40, 0.18) | 0.47 | -0.12 (-0.39, 0.16) | 0.40 |
| RWG between 2 and 4 years | 1,065 | 0.12 (-0.11, 0.34) | 0.31 | 0.21 (-0.04, 0.45) | 0.10 | -0.01 (-0.31, 0.29) | 0.95 | -0.07 (-0.35, 0.22) | 0.65 |
| RWG between 4 and 7 years | 1,168 | -0.10 (-0.32, 0.13) | 0.37 | -0.08 (-0.33, 0.16) | 0.50 | 0.14 (-0.16, 0.43) | 0.36 | 0.11 (-0.18, 0.39) | 0.46 |
| RWG between 7 and 15 years | 1,161 | 0.15 (-0.06, 0.36) | 0.16 | 0.02 (-0.22, 0.25) | 0.88 | 0.21 (-0.08, 0.49) | 0.16 | 0.40 (0.13, 0.68) | <0.001 |
| *Linear growth* | | | | | | | | | |
| CLG between 2 and 4 years | 1,085 | 0.09 (-0.13, 0.31) | 0.44 | 0.15 (-0.10, 0.39) | 0.24 | -0.20 (-0.50, 0.10) | 0.18 | -0.18 (-0.46, 0.10) | 0.21 |
| CLG between 4 and 7 years | 1,204 | 0.03 (-0.17, 0.23) | 0.77 | -0.03 (-0.25, 0.19) | 0.78 | 0.07 (-0.19, 0.34) | 0.60 | -0.04 (-0.29, 0.22) | 0.77 |
| CLG between 7 and 15 years | 1,174 | 0.04 (-0.16, 0.25) | 0.67 | -0.01 (-0.24, 0.21) | 0.90 | -0.07 (-0.34, 0.21) | 0.64 | 0.16 (-0.11, 0.43) | 0.24 |

Adjusted for age in months at 53 years and sex.

RWG: Relative weight gain i.e. standardised residuals from regression of present weight z-score on previous weight and height z-scores and present height z-scores

CLG: Conditional linear growth i.e. Standardised residuals from regression of present height z-score on previous height and weight z-scores

**Supplementary table 4.** DNAm Age Acceleration at 53 years and pubertal timing adjusted for estimated cell composition

| | | AgeAccelHannum | | AgeAccelHorvath | | AgeAccelLevine | | AgeAccelGrim | |
|---|---|---|---|---|---|---|---|---|---|
| | N | Coefficient (95% CI) | P value | Coefficient (95% CI) | P value | Coefficient (95% CI) | P value | Coefficient (95% CI) | P value |
| *Women* | | | | | | | | | |
| Age at menarche (years) | 617 | -0.01 (-0.23, 0.20) | 0.92 | 0.003 (-0.240, 0.245) | 0.98 | -0.23 (-0.53, 0.07) | 0.13 | 0.07 (-0.22, 0.35) | 0.66 |
| Age at menarche | | | | | | | | | |
| <12 years | 96 | Ref. | | Ref. | | Ref. | | Ref. | |
| ≥12 years | 521 | 0.02 (-0.73, 0.76) | 0.97 | 0.16 (-0.68, 1.00) | 0.70 | -0.04 (-1.09, 1.00) | 0.94 | 0.53 (-0.46,1.53) | 0.29 |
| *Men: Pubertal status at 14-15 years* | | | | | | | | | |
| Fully mature* | 172 | Ref. | | Ref. | | Ref. | | Ref. | |
| Advanced puberty | 211 | 0.14 (-0.62, 0.89) | 0.27** | -0.40 (-1.22, 0.42) | 0.80** | 0.93 (-0.05, 1.90) | 0.05** | -0.31 (-1.26, 0.64) | 0.78** |
| Early puberty | 200 | 0.62 (-0.14, 1.39) | | -0.13 (-0.97, 0.70) | | 0.82 (-0.17, 1.81) | | 0.15 (-0.82, 1.12) | |
| Pre-pubertal | 73 | -0.18 (-1.21, 0.84) | | -0.28 (-1.40, 0.84) | | -0.52 (-1.84, 0.81) | | -0.18 (-1.48, 1.11) | |
| Fully mature | 172 | Ref. | | Ref. | | Ref. | | Ref. | |

| | | | | | | | | | |
|---|---|---|---|---|---|---|---|---|---|
| Later puberty | 484 | 0.29 (-0.36, 0.94) | 0.38 | -0.27 (-0.98, 0.44) | 0.45 | 0.67 (-0.18, 1.51) | 0.12 | -0.10 (-0.92, 0.72) | 0.81 |

Adjusted for age in months at 53 years. *Fully mature group are the group who experienced the earliest pubertal timing. **p-value from lrtest comparing models with and without categorical puberty variable

**Supplementary table 5.** DNAm Age Acceleration at 60-64 years and conditional growth

| | | AgeAccelHannum | | AgeAccelHorvath | | AgeAccelLevine | | AgeAccelGrim | |
|---|---|---|---|---|---|---|---|---|---|
| | N | Coefficient (95% CI) | P value | Coefficient (95% CI) | P value | Coefficient (95% CI) | P value | Coefficient (95% CI) | P value |
| *Relative weight gain* | | | | | | | | | |
| RWG between birth and 2 years | 402 | -0.11 (-0.55, 0.33) | 0.61 | -0.11 (-0.58, 0.36) | 0.64 | -0.21 (-0.83, 0.41) | 0.51 | -0.50 (-0.95, -0.05) | 0.03 |
| RWG between 2 and 4 years | 385 | 0.51 (0.08, 0.95) | 0.02 | 0.59 (0.13, 1.06) | 0.01 | 0.54 (-0.07, 0.40) | 0.08 | 0.05 (-0.41, 0.50) | 0.84 |
| RWG between 4 and 7 years | 441 | -0.39 (-0.80, 0.03) | 0.07 | -0.12 (-0.57, 0.33) | 0.61 | -0.20 (-0.79, 0.40) | 0.52 | -0.35 (-0.79, 0.10) | 0.13 |
| RWG between 7 and 15 years | 450 | 0.17 (-0.19, 0.54) | 0.35 | 0.03 (-0.39, 0.45) | 0.89 | 0.69 (0.12, 1.26) | 0.02 | 0.38 (-0.04, 0.79) | 0.08 |
| *Linear growth* | | | | | | | | | |
| CLG between 2 and 4 years | 393 | -0.16 (-0.59, 0.26) | 0.45 | 0.21 (-0.25, 0.66) | 0.37 | -0.37 (-0.97, 0.22) | 0.22 | -0.10 (-0.54, 0.34) | 0.66 |
| CLG between 4 and 7 years | 441 | -0.11 (-0.48, 0.26) | 0.55 | -0.27 (-0.66, 0.13) | 0.19 | -0.17 (-0.69, 0.36) | 0.54 | -0.07 (-0.47, 0.32) | 0.71 |

| | | | | | | | | | |
|---|---|---|---|---|---|---|---|---|---|
| CLG between 7 and 15 years | 454 | 0.14 (-0.23, 0.50) | 0.46 | 0.51 (0.09, 0.93) | 0.02 | 0.72 (0.16, 1.29) | 0.01 | 0.26 (-0.16, 0.67) | 0.23 |

Adjusted for age in months at 60-64 years and sex.

RWG: Relative weight gain i.e. standardised residuals from regression of present weight z-score on previous weight and height z-scores and present height z-score

CLG: Conditional linear growth i.e. Standardised residuals from regression of present height z-score on previous height and weight z-scores

**Supplementary table 6.** DNAm Age Acceleration at 60-64 years and pubertal timing

| | | AgeAccelHannum | | AgeAccelHorvath | | AgeAccelLevine | | AgeAccelGrim | |
|---|---|---|---|---|---|---|---|---|---|
| | N | Coefficient (95% CI) | P value | Coefficient (95% CI) | P value | Coefficient (95% CI) | P value | Coefficient (95% CI) | P value |
| *Women* | | | | | | | | | |
| Age at menarche (years) | 212 | 0.26 (-0.14, 0.66) | 0.19 | 0.55 (0.13, 0.97) | 0.01 | 0.653 (0.002, 1.305) | 0.049 | 0.38 (-0.09, 0.85) | 0.11 |
| Age at menarche | | | | | | | | | |
| <12 years | 34 | | | | | | | | |
| ≥12 years | 178 | 1.34 (-0.02, 2.70) | 0.05 | 1.09 (-0.36, 2.55) | 0.14 | 2.13 (-0.09, 4.36) | 0.06 | 1.50 (-0.09,3.10) | 0.06 |
| *Men: Pubertal status at 14-15 years* | | | | | | | | | |
| Fully mature* | 73 | Ref. | | Ref. | | Ref. | | Ref. | |
| Advanced puberty | 92 | -0.70 (-2.11, 0.71) | 0.07** | -0.44 (-1.96, 1.08) | 0.28** | -0.70 (-2.54, 1.14) | 0.62** | -1.29 (-2.68, 0.09) | 0.10** |
| Early puberty | 81 | -0.41 (-1.86, 1.04) | | -1.06 (-2.62, 0.50) | | -1.65 (-3.54, 0.25) | | -1.23 (-2.66, 0.20) | |
| Pre-pubertal | 24 | -1.76 (-3.88, 0.35) | | -1.80 (-4.08, 0.49) | | -1.85 (-4.62, 0.92) | | -1.29 (-3.38, 0.79) | |
| Fully mature | 73 | Ref. | | Ref. | | Ref. | | Ref. | |
| Later puberty | 197 | -0.72 (-1.95, 0.51) | 0.25 | -0.86 (-2.19, 0.46) | 0.20 | -1.23 (-2.84, 0.37) | 0.13 | -1.27 (-2.47, -0.06) | 0.04 |

Adjusted for age in months at 60-64 years. *Fully mature group are the group who experienced the earliest pubertal timing. **p-value from lrtest comparing models with and without categorical puberty variable
